# Humoral immune response in inactivated SARS-CoV-2 vaccine: When should a booster dose be administered?

**DOI:** 10.1101/2021.07.08.21260194

**Authors:** Ertan Kara, Ferdi Tanir, Hakan Demirhindi, Burak Mete, Filiz Kibar, Salih Cetiner, Aslihan Candevir, Ayşe Inaltekin

## Abstract

**Background:** For a sustained and essential protective antibody response, it is important to understand how long the humoral immune response induced by the SARS-CoV-2 inactivated vaccine persists.

**Aims:** This study aimed to detect the first and third-month concentrations and seroconversion rates of the antibodies induced by the inactivated vaccine.

**Study Design:** This is a vaccine efficacy study.

**Methods:** The study included 272 health workers who were vaccinated at days 0 and 28 by the inactivated SARS-CoV-2 vaccine (3μg/0.5ml). Anti-S-RBD-IgG and total anti-spike/anti-nucleocapsid-IgG antibody concentrations and seroconversion rates were examined in vaccinated health workers at the 1^st^ and 3^rd^ months after the vaccination. The test method used for the qualitative detection and differentiation of IgG antibodies (indirect method) to SARS-CoV-2 is a chemiluminescence reaction (CLIA).

**Results:** The mean age of the health workers was 38.93±10.59 (min:21-max:64). A total of 45(16.5%) participants declared to have had COVID-19 before the first dose of the inactivated vaccine. The participants were found to be reactive for anti-S-RBD-IgG antibodies by 98.2% and 97.8% at the first and third months, respectively, after the administration of the second dose. The decrease in the mean plasma concentrations of anti-S-RBD IgG was observed as 56.7% in the cohort with only two doses of the vaccine (1^st^ month:42.4AU/ml versus 3^rd^ month: 18.2AU/ml). In the cohort with a history of COVID-19 prior to the vaccination, the decrease was observed as 25.1% (1^st^ month:58.29 versus 3^rd^ month:43.64 AU/ml) and at a mean of 57.4 (0-90) days prior to vaccination, the decrease was of 43.1% (1^st^ month:55.05 AU/ml versus 3^rd^ month:31.28 AU/ml), keeping more stable in participants infected at a mean of 183.1 (91-330) days prior to vaccination (a decrease of 5.2%; with 62.34 AU/ml at 1^st^ and 59.08 AU/ml at 3^rd^ months). Anti-S-RBD concentrations were observed to increase 10-fold (30.44 AU/ml at 1^st^ and 310.64 AU/ml at 3^rd^ months) in participants infected after the vaccination and to decrease among people aged 50 years and older.

**Conclusion:** Antibody concentrations at the 1^st^ and 3^rd^ months after the vaccination with two doses of the inactivated SARS-CoV-2 vaccine were found to be decreased, but still detectable (except in one participant). As participants who had COVID-19 at a mean of 181 (90-330) days before the vaccination presented with a more stable antibody level, it can be concluded that a booster at months 6-12, resulting in a schedule of 0-1-6 months, is recommended for the inactive SARS-CoV-2 vaccination.

## Introduction

The World Health Organization declared the outbreak of coronavirus disease in 2019 (COVID-19) to be a public health emergency of international concern on 30 January 2020, and a pandemic on 11 March 2020. Nearly 80% of COVID-19 patients were reported to have mild-to-moderate symptoms, while nearly 20% were reported to have developed serious manifestations such as severe pneumonia, acute respiratory distress syndrome (ARDS), sepsis and even death [1]. The number of COVID-19 cases has increased at an astonishing rate worldwide. Severe acute respiratory syndrome–coronavirus (SARS-CoV)-2 (SARS-CoV-2), the causative virus of the ongoing pandemic, belongs to the genus *Betacoronavirus* (β-CoV) of the family *Coronaviridae* [2]. SARS-CoV-2, SARS-CoV and the Middle Eastern respiratory syndrome-related coronavirus (MERS-CoV) constitute the three most life-threatening species among all human coronaviruses. SARS-CoV-2 possesses a linear single-stranded positive sense RNA genome, encoding 4 structural proteins [spike (S), envelope (E), membrane (M), and nucleocapsid (N)] of which S is a major protective antigen that elicits highly potent neutralising antibodies (NAbs), 16 non-structural proteins (nsp1-nsp16) and several accessory proteins [3]. Purified inactivated viruses have been traditionally used for vaccine development and such vaccines have been found to be safe and effective for the prevention of diseases caused by viruses like *Influenza virus* and *Poliovirus* [4, 5]. A vaccine against SARS-CoV-2 might act against infection, disease, or transmission, and a vaccine capable of reducing any of these elements could contribute to disease control. However, the most important efficacy endpoint, i.e. protection against severe disease and death, is difficult to assess in phase 3 clinical trials [6]. Reinfections by seasonal coronaviruses occur 6-12 months after the previous infection, indicating that protective immunity against these viruses may be short-lived [7]. Early reports documenting rapidly declining antibody titres in convalescent SARS-CoV-2 patients in the first several months after infection suggested that protective immunity against SARS-CoV-2 might be similarly transient [8-10]. It was also suggested that SARS-CoV-2 infection might fail to elicit a functional germinal centre response, which would interfere with the generation of long-lived plasma cells [11]. Later reports analysing samples collected approximately 4 to 6 months after infection indicated that SARS-CoV-2 antibody titres declined more slowly [12]. Durable serum antibody titres are maintained by long-lived plasma cells, which are non-replicating, antigen-specific plasma cells detected in bone marrow long after the disappearance of the antigen [13]. It was reported that a rapid decrease in antibody titrations occurred in a few months after natural infection with SARS-CoV-2 and that humoral immunity might be short-lived [11, 13]. How long the antibody response elicited by the inactivated vaccine can be measured, and what the antibody levels are in people who have been infected before and after the vaccination are questions that need to be answered. The answers to these questions will contribute to the determination of a public vaccination schedule especially for risk groups. This study aimed to detect the change in antibody levels in the first and third months after the administration of two doses of inactivated SARS-CoV-2 vaccine, and a possible need for a booster dose.

## MATERIAL AND METHODS

### Study design and participants

This study constituted the second step of a vaccine efficacy project carried at Cukurova University in May 2021 and included health workers who had been vaccinated with two doses of the inactivated SARS-CoV-2 vaccine in the context of a public vaccination program by the Turkish Ministry of Health, with health workers constituting the priority group in the vaccination target population. It was not a trial study as the participants had already been vaccinated before the study. The incidence of adverse events in the vaccine cohort was determined to be 19% after the administration of 3 μg (corresponding to 600 SU) of the inactivated SARS-CoV-2 vaccine during Phase 2 clinical trials [14]. The minimum sample size was calculated as 220 participants by assuming type-1 error as 0.05 and type-2 error as 0.20 considering reported adverse events rates. The participants were randomly selected from a list of 3,000 health workers. Two substitution lists were also prepared by randomisation. The total number of participants was 282 health workers at the first step of the study performed at the end of the first month after vaccination, but this number decreased to 272 at this second step of the study performed at the end of the third month after the second dose of the vaccine when ten participants resigned from the study of their own free will. Blood samples were taken after the participants had been informed about the study and had signed an informed consent form. They answered a questionnaire form about sociodemographic characteristics, history of natural COVID-19 infection, vaccination and adverse events.

### Vaccine information

The generic name of the vaccine administered to health workers by the Turkish Ministry of Health is “inactivated SARS-CoV-2 vaccine (Vero Cell) (CoronaVac™)”, with aluminium hydroxide developed by Sinovac Biotech Ltd., Life Sciences Lab., China. The vaccine was administered intramuscularly in the deltoid region of the upper arm with a dosage of 3 μg/0.5 ml. The two doses were administered 28 days apart. Vaccines were transferred, stored and administered following cold chain principles already in use by and therefore familiar to health institutions performing vaccinations.

### Immunogenicity assessments

The project aimed to measure anti-SARS-CoV-2 S-RBD (anti-S-RBD) immunoglobulin G (IgG), total anti-spike and anti-nucleocapsid immunoglobulin IgG antibody concentrations in the same individuals repetitively in the first, third and sixth months. This second step of the project aimed to reveal anti-S-RBD-IgG, total anti-spike/anti-nucleocapsid IgG concentrations at the end of the third month after the administration of the second dose of the inactivated vaccine. The third step of the project has been planned to be performed in the sixth month after the second dose. About 5 ml of blood samples were collected into biochemistry tubes with vacuum gel. The sera were extracted by centrifugation at 3000 gv for 10 minutes and kept at 2-8°C for 1-3 days. Test calibrators and controls were performed first. After the control results were observed to be within the expected ranges, the samples were tested by trained experts in the accredited (by the Joint Commission International (JCI) since 2006) Central Laboratory of Cukurova University Balcali Hospital.

The blood samples stored at 2-8°C were brought to room temperature on the day the tests were performed and examined collectively after all participants were sampled. The participants were not informed about their antibody concentration at the first month (in a single-blind format) to avoid a possible effect on the declaration of clinical complaints or outcomes as they would be followed repetitively for 6 months. The test method used for the qualitative detection and differentiation of IgG antibodies (indirect method) to SARS-CoV-2 in human serum and serum in separating gel tubes is a chemiluminescence immunoassay (CLIA) reaction based on the measurement of light signals by a photomultiplier as relative light units (RLUs) proportional to the concentration of antibodies present in the sample [15, 16]. The test is only for use according to the Food and Drug Administration’s Emergency Use Authorization [17]. The SARS-CoV-2 S-RBD IgG test is also an indirect CLIA and has a high correlation with VNT50 titres (R=0.712), where VNT stands for “Virus Neutralization Test” which is a gold standard for quantifying the titre of neutralising antibodies (nAbs) for a virus [18]. The results are expressed in absorbance units (AU/mL) and reported to the end-user as “Reactive (i.e. a result ≥1.00 AU/mL)” or “Non-Reactive (i.e. a result <1.00 AU/mL)” [16].

### Statistical analyses

Data were examined using the SPSS 22 statistical analyses package (2013, IBM, New York, U.S.A). Following normality testing (Kolmogorov-Smirnov), data were analysed by Wilcoxon and Mc Nemar tests. A value of p <0.05 was considered significant.

## Results

After the resignation of ten participants of their own will following the first step of the project, remaining 272 health workers participated in the second step performed after three months following the second dose of the inactivated vaccine. The mean age of the participants was 38.93±10.59 (min:21-max:64). A total of 45(16.5%) participants declared to have had COVID-19 before the administration of the first dose of the inactivated vaccine, while nobody in the study cohort was infected with COVID-19 between the first and second dose of the vaccine. The mean time interval between the disease and the administration of the first dose of the vaccine was calculated as 111 days (min 29; max 330 days). After the second dose 5(1.8%) participants declared to have been infected with COVID-19, but all with mild symptoms. These cases were confirmed through the health registry system and the mean infection duration was found to be 60.00±12.24 days (min:40-max:70). The socio-demographic characteristics and antibody concentrations of the participants at the first and third months are presented in table 1 (Table.1)

**Table 1.**
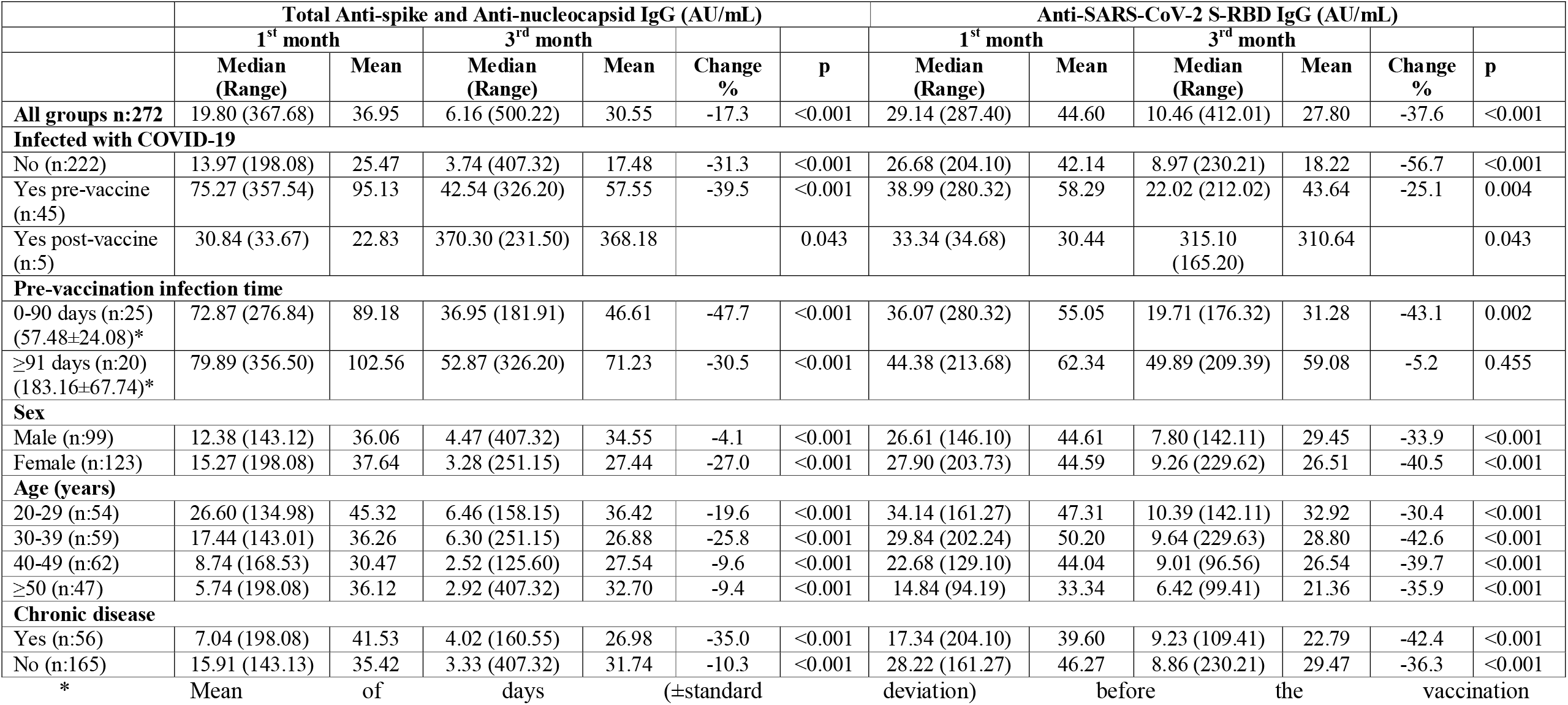
Antibody concentrations at the end of the first and third months after the administration of the second dose of the inactivated vaccine.

The first and third months’ distributions of anti-S-RBD IgG (Fig.1) and total anti-spike/anti-nucleocapsid IgG (Fig.2) concentrations of participants with the history of natural COVID-19 infection (pre-vaccination or post-vaccination) and of those vaccinated with two doses without any history of COVID-19 are presented in figures 1 and 2 (Fig.1-2).

**Figure 1.**
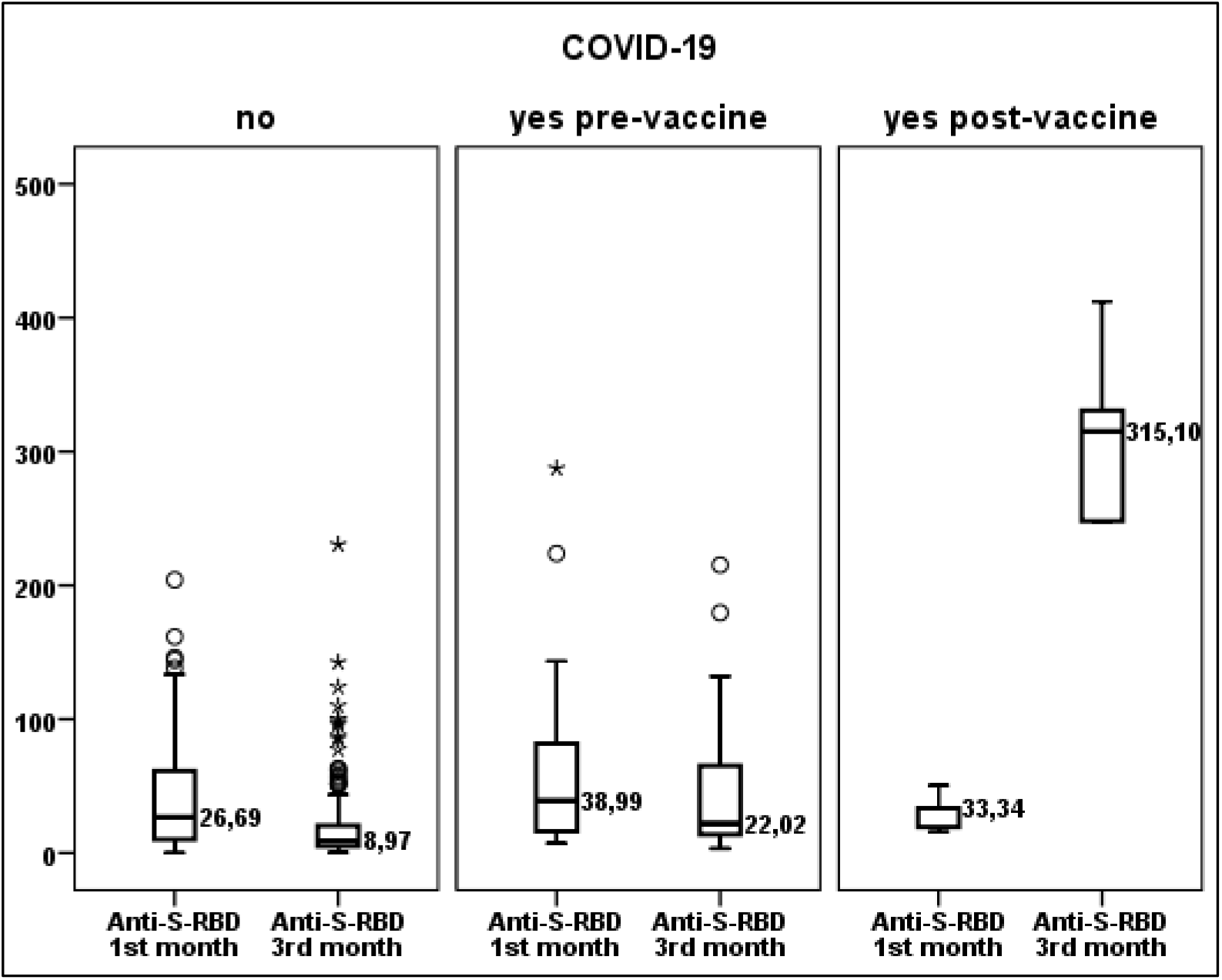
Anti-S-RBD IgG concentrations according to the history of natural COVID-19 infection in the first and third months after the vaccination

**Figure 2.**
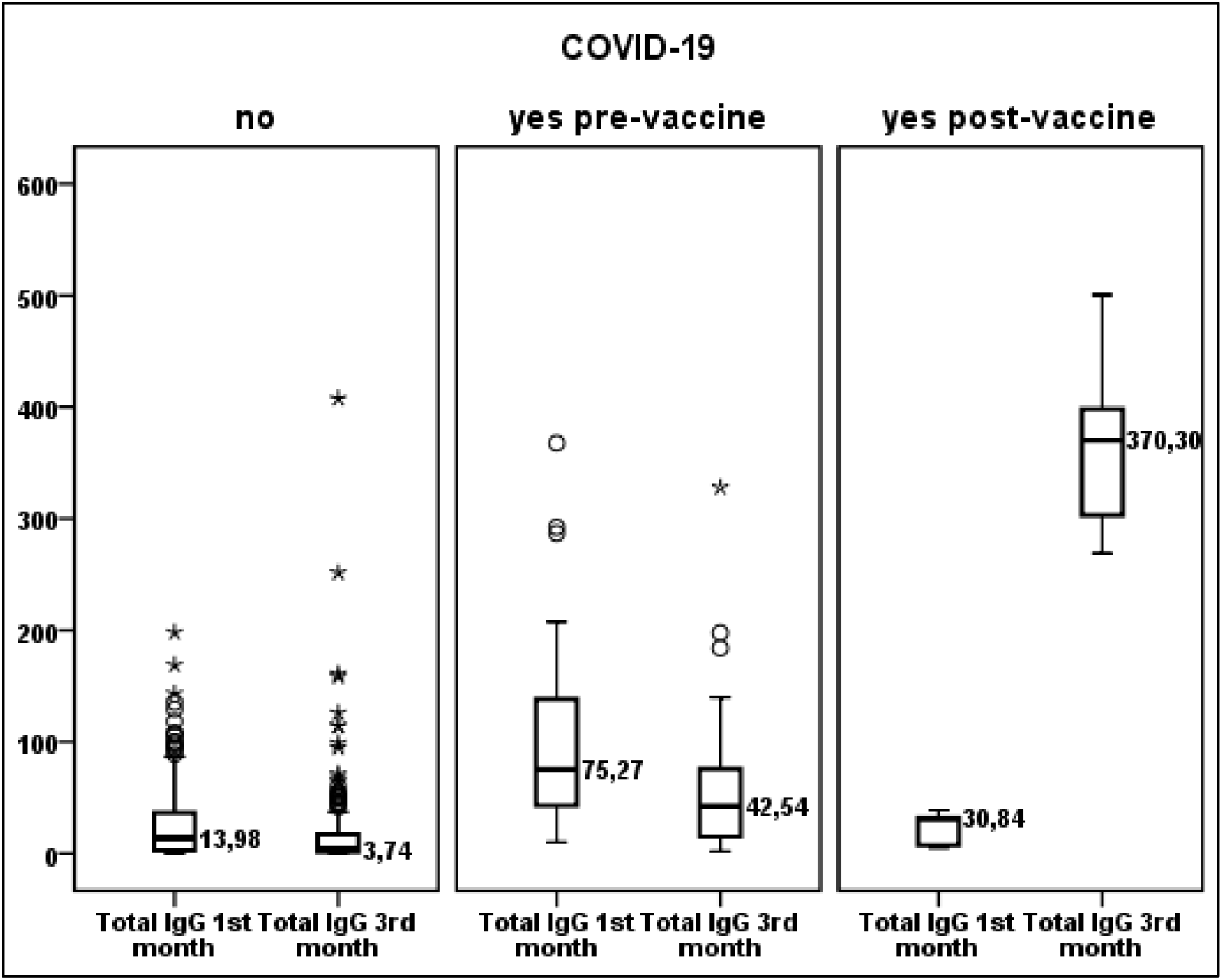
Total anti-spike/anti-nucleocapsid IgG concentrations according to the history of natural COVID-19 infection in the first and third months after the vaccination

When the antibody concentrations at the first and third months were compared in the total group and subgroups, it was assessed that the antibody concentrations were statistically significantly decreased at the third month, but with the median concentrations never falling below the reactive level. The exception was in the subgroup naturally infected with COVID-19 nearly six months (mean 183.16 (range 90-300) days) before the start of the vaccination, where the decrease in the antibody concentrations was not found to be statistically significant (Table 1, Fig.3).

**Figure 3.**
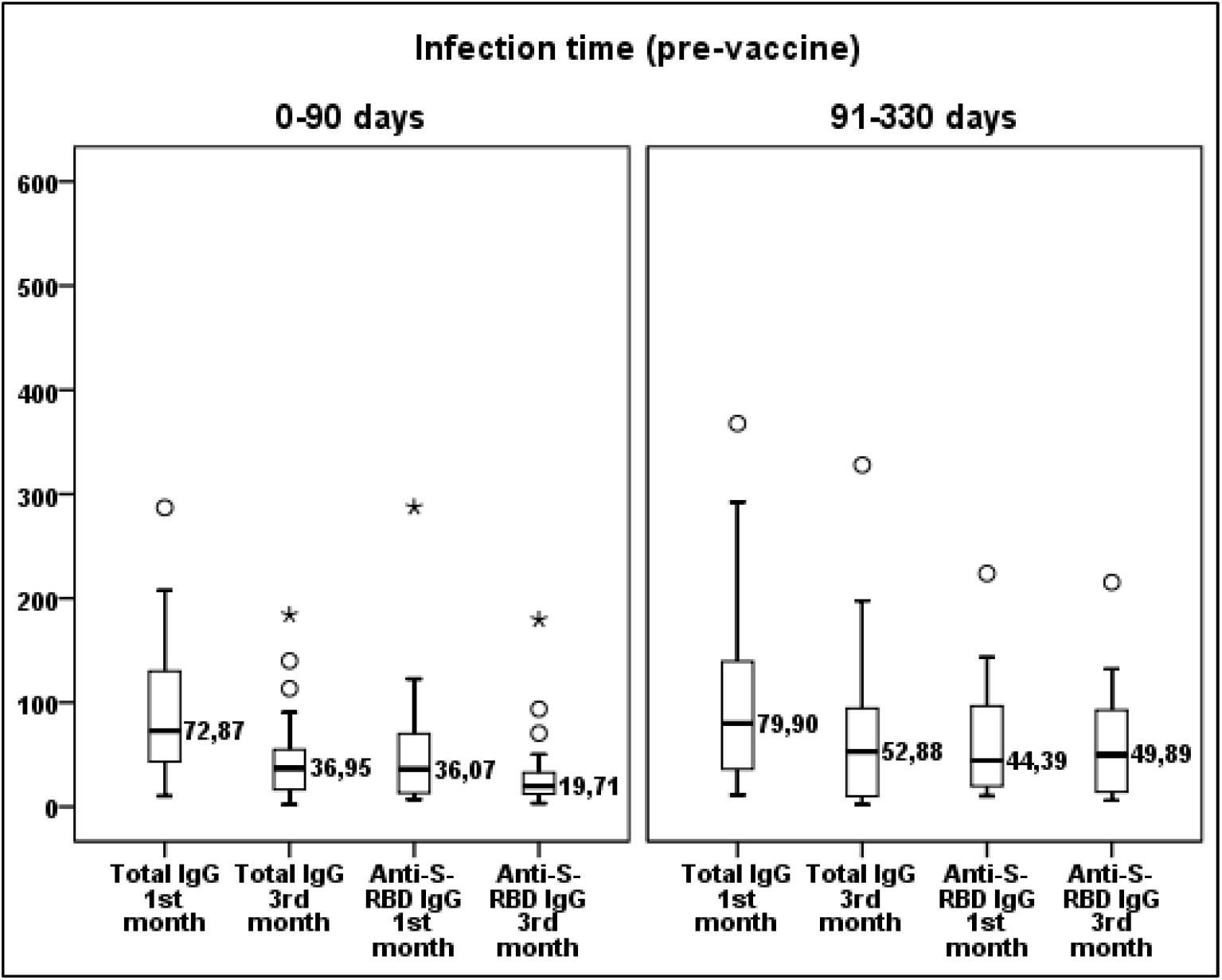
Total anti-spike/anti-nucleocapsid IgG and total anti-S-RBD IgG concentrations according to the time of having natural COVID-19 infection before the vaccination

After the administration of the second dose of the inactivated vaccine, 98.2% of participants were found to be reactive for anti-S-RBD-IgG antibodies at the first month, and %97.8 at the third month, while for total anti-spike/anti-nucleocapsid-IgG antibodies 93.0% at the first month and 87.5% at the third month. When the first and third months’ results were compared, the decrease in the seroconversion rates for anti-S-RBD-IgG antibodies was not statistically significant (Table 2).

**Table 2.** Seroconversion rates at the end of the first and third months after the second dose of vaccination with the inactivated vaccine.

|  | Total Anti-spike/Anti-nucleocapsid IgG |  | p | Anti-SARS-CoV-2 S-RBD IgG |  | p |
| --- | --- | --- | --- | --- | --- | --- |
|  | 1<sup>st</sup> month | 3<sup>rd</sup> month |  | 1<sup>st</sup> month | 3<sup>rd</sup> month |  |
|  | N/R* (%) | N/R* (%) |  | N/R* (%) | N/R* (%) |  |
| **All groups n:272** | 7.0/93.0 | 12.5/87.5 | **0.004** | 1.8/98.2 | 2.2/97.8 | 1.000 |
| **Infected with COVID-19** |  |  |  |  |  |  |
| No (n:222) | 8.6/91.4 | 15.3/84.7 | **0.004** | 2.3/97.7 | 2.7/97.3 | 1.000 |
| Yes pre-vaccine (n:45) | 0.0/100.0 | 0.0/100.0 | 1.000 | 0.0/100.0 | 0.0/100.0 | 1.000 |
| Yes post-vaccine (n:5) | 0.0/100.0 | 0.0/100.0 | 1.000 | 0.0/100.0 | 0.0/100.0 | 1.000 |
| **Pre-vaccine infection time** |  |  |  |  |  |  |
| 0-90 days (n:25) | 0.0/100.0 | 0.0/100.0 | 1.000 | 0.0/100.0 | 0.0/100.0 | 1.000 |
| ≥91 days (n:20) | 0.0/100.0 | 0.0/100.0 | 1.000 | 0.0/100.0 | 0.0/100.0 | 1.000 |
| **Sex** |  |  |  |  |  |  |
| Male (n:119) | 8.4/91.6 | 15.1/84.9 | **0.039** | 2.5/97.5 | 4.2/95.8 | 0.500 |
| Female (n:153) | 5.9/94.1 | 10.5/89.5 | 0.092 | 1.3/98.7 | 0.7/99.3 | 1.000 |
| **Age (years)** |  |  |  |  |  |  |
| 20-29 (n:68) | 2.9/97.1 | 8.8/91.2 | 0.219 | 2.9/97.1 | 1.5/98.5 | 1.000 |
| 30-39 (n:79) | 5.1/94.9 | 15.2/84.8 | 0.008 | 0.0/100.0 | 2.5/97.5 | 0.500 |
| 40-49 (n:73) | 9.6/90.4 | 11.0/89.0 | 1.000 | 1.4/98.6 | 0.0/100.0 | 1.000 |
| ≥50 (n:52) | 11.5/88.5 | 15.4/84.6 | 0.625 | 3.8/96.2 | 5.8/94.2 | 1.000 |
| **Chronic disease** |  |  |  |  |  |  |
| Yes (n:67) | 10.4/89.6 | 10.4/89.6 | 1.000 | 4.5/95.5 | 4.5/95.5 | 1.000 |
| No (n:204) | 5.9/94.1 | 13.2/86.8 | **<0.001** | 1.0/99.0 | 1.5/98.5 | 1.000 |

\*N:Non-reactive, R:Reactive, statistically significant values in bold

Some adverse reactions were reported by 10% of participants at the end of the third month after the second dose of the inactivated vaccine. The most persisting adverse reactions were pain at the injection site, back pain, joint pain and headache. No neurological adverse reactions were observed.

## Discussion

Infection or vaccination induces the development of long-lived plasma cells in the bone marrow (BMPCs) to provide a persistent and essential source of protective antibodies [19]. It is still not known whether these BMPCs are induced or not in patients infected with SARS-CoV-2. Some recent studies examining antigen-specific serum antibodies against SARS-CoV-2 in convalescent patients reported a rapid decline in these concentrations, raising concerns about the short longevity of humoral immunity against the SARS-CoV-2 virus [13, 20, 21]. This fact is similarly observed in cases of infection with other human coronaviruses, where antibodies decrease over time leading to reinfection with homologous viruses after as short a time as 80 days [22]. Information about humoral immunity after a natural infection has begun to emerge, but there is no information yet about vaccine-induced humoral immunity. Our study aimed to answer the question of how long the plasma life of antibodies induced by inactivated SAR-CoV-2 vaccine is. In our study cohort, plasma anti-S-RBD IgG and total anti-spike/anti-nucleocapsid IgG concentrations were examined comparing three subgroups: between the first and third months after the administration of the inactive vaccine, the mean anti-S-RBD IgG concentrations were found to have decreased by 56.7% in participants without any history of COVID-19 infection who had received two-doses (0-28-day regime) of the vaccine, while by 25.1% among those having a history of natural COVID-19 infection prior to the vaccination. Further analyses indicated that when the interval between the natural COVID-19 infection and the first dose of the vaccination was between 0 and 90 days, the decrease in anti-S-RBD concentrations was observed to be 43.1%, while to be 5.2% when the interval was between 91 and 330 days. The antibody levels presented a stable course in the 91-330 days cohort. A 10-fold increase in the mean anti-S-RBD concentrations was observed in participants who had been infected after the start of the vaccination. Similar decreases in total anti-spike/anti-nucleocapsid IgG levels were found among participants with no history of COVID-19 infection and similarly again a 16-fold increase was observed in case of infection after the vaccination.

Turner et al. collected blood samples of 77 patients (aged 21-69 years), who had recovered from a mild COVID-19 infection, at the 1st, 4th, 7th and 11th months after the onset of symptoms. Bone marrow aspirates from 18 of these patients were also collected at 7th and 8th months. They reported that the anti-SARS-CoV-2 spike (anti-S) IgG antibodies were detectable for 11 months after the onset of symptoms, with a rapid decrease at the first 4 months (estimated half-life of 116.5 days) and a slower decay rate resulting in a more stable phase (estimated half-life of 686.3 days) in the following 7 months. The titres were found to be correlated with the frequency of S-specific BMPCs obtained from bone marrow aspirates of convalescent patients. Neither anti-S antibodies, nor BMPCs were detectable in 11 healthy volunteers (aged 23–60) with no history of SARS-CoV-2 infection. As S-binding BMPCs were observed to be quiescent, Turner et al. reported that they might belong to a long-lived compartment in the marrow. Circulating resting memory B cells directed against the S protein were detected in the convalescent individuals. They concluded that SARS-CoV-2 infection induced a robust antigen-specific, long-lived humoral immune response in humans [23]. The results of our study showed a rapid decrease in the anti-S-RBD IgG concentrations induced by the inactivated SARS-CoV-2 vaccine between 1^st^ and 3^rd^ months in people who have never had the disease. In addition, when vaccinated subjects with a history of COVID-19 were compared, more stable antibody concentrations were observed in subjects who were infected nearly 6 months (mean of 183.6 (range 90-330) days) before the first dose of vaccination. Gaebler et al. reported a significant decrease in the titres of anti-S-RBD IgM and IgG antibodies at 1.3 and 6.2 months after infection with SARS-CoV-2 in 87 patients, in contrast to the number of RBD-specific memory B cells, which remained unchanged at 6.2 months after infection [24]. Memory cells remain silent until they encounter the antigen again and produce a much faster and more powerful antibody response on the next encounter with the same antigen [25].

In our study, we observed a 10-fold increase in the antibody concentrations of the subjects who were not infected before the vaccination, but were infected after the vaccination (five health workers). This severe increase in antibody concentration suggests a secondary immune response. Based on this, the inactivated SARS-CoV-2 vaccine could be assumed to induce the production of memory-cells. The early and mid-term results of our study showed that the humoral immunity induced by inactivated SARS-CoV-2 vaccine was similar to the humoral immune response induced by natural infection. Higher and stable antibody concentrations in participants who had been infected an average of 183.16±67.74 (90-330) days before the vaccination, indicate the need for a booster dose resulting in a final schedule of 0-28-180 days (i.e. 0-1-6 months). The range for the 3^rd^ dose can also be suggested as 6-11 months in order to induce a stronger and longer-term protective humoral immunity.

On the other hand, low antibody concentrations in people over 50 years of age, with weak/absent immune response or with chronic disease, indicate that seroconversion will decrease to an undetectable level in an earlier period. A safe, effective and ethical herd immunity can be achieved through controlled vaccination programs. Booster vaccines and updated vaccines will almost certainly be required to maintain population-level herd immunity with diminishing humoral immunity against an extremely diffuse and contagious virus and to maintain immunity against re-infection by SARS-CoV-2 and its inevitable genetic variants [26]. It is of utmost importance to understand the efficacy process of vaccine-induced immunity in order to develop effective vaccination programs and vaccination schedules.

## Data Availability

This is a vaccine efficacy study and not a clinical study. Data available on request from the authors.

## Ethical considerations

The study was approved by (1)The Scientific Research Platform of the Turkish Ministry of Health and (2) the Scientific Ethics Committee for Non-interventional Clinical Studies of the Cukurova University Faculty of Medicine (Adana, Turkey) (Decree no. 36, Meeting No. 108, 12.02.2021). The official invitation letters with the list of the randomly selected participants and substitutions were sent to the department headships in order to let them invite the selected staff to participate in the study. Informed written consents were obtained from all participants after required acknowledgement.

## Conflict of interest

The authors declare that they have no known competing financial interests or personal relationships that could have appeared to influence the work reported in this paper.

All of the authors declare that they have all participated in the design, execution, and analysis of the paper and that they have approved the final version in accordance to the ICJME recommendations.

The submitted paper has been shared in a non-profit preprint server for health sciences, but it is not under publication or consideration for publication elsewhere.

